# Features of C-reactive protein in COVID-19 patients within various period: a cohort study

**DOI:** 10.1101/2020.10.26.20219360

**Authors:** Guoxin Huang, Gaojing Qu, Hui Yu, Meiling Zhang, Xiaoming Song, Lei Chen, Haoming Zhu, Yunfu Wang, Bin Pei

## Abstract

**BACKGROUND:** Coronavirus disease 2019 (COVID-19) has been declared as a threat to the global. Due to the lack of efficient treatments, indicators were urgently needed during the evolvement of disease to analyze the illness and prognosis and prevent the aggravation of COVID-19.

**METHODS:** All laboratory confirmed COVID-19 patients hospitalized in Xiangyang No.1 People’s Hospital were included. Patients’ general information, clinical type, CRP value and outcome were collected. CRP values of all patients during disease course from different initial time were analyzed.

**RESULTS:** The 131 enrolled patients were 50.13±17.13 years old. All cases underwent 724 tests of CRP since symptom onset, 53.18% of the test results were abnormal and the median value was 9.52(2.63-34.10) mg/L. The first median value on the day 8 from exposure onset was 39.08(11.92-47.89) mg/L then fluctuated around it until the day 28. The CRP median increased from 15.93 mg/L to 41.44 mg/L and then decreased to 18.26 mg/L before transformation of severe type, and then increased to 62.25 mg/L on the transforming date. Conversely, the CRP median increased from 56.17 mg/L 102.75 mg/L before transformation of critical type but decreased to 68.68 mg/L on the transforming date. The changes of CRP median over time before death ranged from 77.77 mg/L to 133.52 mg/L.

**CONCLUSIONS:** CRP increased before symptom onset and substantially increased during the early-to-mid stage (especially early stage), which was different from other virus-infected diseases. The changes of CRP before the transformation of clinical type was inconsistent with the aggravating of illness. And the CRP maintained over 100.00 mg/L prompted poor prognosis.

## 1. Introduction

Coronavirus disease 2019 (COVID-19) was caused by severe acute respiratory syndrome coronavirus 2 (SARS-CoV-2) (1, 2). The World Health Organization (WHO) declared COVID-19 as a global pandemic on Mar 11, 2020 (3). C-reactive protein (CRP) was reported in 1930 by Tibet and Francis firstly (4). Several cross-section researches in the early stage of COVID-19 outbreak indicated that CRP was abnormal in COVID-19 patients (5, 6). Zhong *et al*. indicated that CRP abnormally increased in 481 out of 793 tests (60.7%) among 99 COVID-19 cases (7).

A study revealed that CRP value was positively correlated with the diameter of lung lesions, and showed significant differences in different clinical type through the analysis of admission results (8, 9). Herold *et al*. collected the admission value and maximum of CRP and IL-6 before intubation. They found that the maximum of IL-6 and CRP value could be used as indexes to predict the need for mechanical ventilation of COVID-19 patients (10). Moreover, laboratory values during pre-partum and post-partum were analyzed to detect the changes of CRP during the pre-partum and post-partum. The results revealed an increased white blood cell (WBC) while decreased lymphocyte (LYMPH) but higher CRP value in COVID-19 confirmed pregnancy than normal pregnancy after partum (11). Researchers reported dynamic profile of WBC count, neutrophil count, and lymphocyte count, etc. of 33 COVID-19 patients in different outcomes since symptom onset, but didn’t study the changes of CRP (12). Similarly, the temporal changes in D-dimer, lymphocyte count, IL-6, serum ferritin, etc. of patients with different outcomes were reported by Zhou *et al*. from disease onset but didn’t contain the changes of CRP (13). Therefore, there lack of systematic research on the changes of CRP with different initial time or during transformed period of severity classification and the relationship between CRP and the evolving illness in COVID-19 patients. In order to reveal the features of CRP during different periods, we included all confirmed COVID-19 patients hospitalized in Xiangyang No.1 People’s hospital, and analyzed all CRP results tested in outpatient and hospitalization.

## 2 Materials and Methods

### 2.1 STUDY DESIGN

This study was a bidirectional observational cohort study. This cohort was established on Feb 9, 2020, all suspected and laboratory-confirmed COVID-19 patients hospitalized in Xiangyang No.1 People’s Hospital affiliated hospital of Hubei University of Medicine before Feb 28, 2020 were included in this cohort. All information was traced back to Jan 23, 2020. The last day of follow-up was Mar 28, 2020. Admission standard and Clinical classifications were made according to the *Diagnosis Guidance for Novel Coronavirus Pneumonia* (14). The study was approved by the ethics review board of Xiangyang No.1 People’s Hospital (No. 2020GCP012) and registered at the Chinese Clinical Trial Registry as ChiCTR2000031088. Informed consent from patients has been exempted since this study does not involve patients’ personal privacy neither incur greater than the minimal risk.

### 2.2 DATA COLLECTION

Data were extracted from the hospital information system by two groups (two researchers per group) and cross-checked individually. Gender, age, all CRP test results, exposure date, disease onset date, transforming date of severe and critical type, outcome, death date, etc. were collected. The data within the course of 1^st^-30th days were statistic analyzed. The distributions of CRP median value in course/period were plotted with an interval of 1 day, 2 days, and 5 days (T1, T2, T3…Tn represented the 5-days unit successively). In this cohort, symptom onset was regarded as disease onset and the corresponding date was set as the 1^st^ day to record data from symptom onset. For patients with clear exposure date and the exposure day was only one day, the exposure date was set as the 1^st^ day to record data when studying the CRP changes from exposure onset. The day when the clinical type changes to severe or critical type was regarded as the “0 day” to study the distribution of CRP before and after the typing transformation. The death date was set as “day -1” and selected the day -1 to day -16 with the tests>4 within two days as the duration to calculate the CRP median value per two days and plot the distribution when studying the CRP features before death. Two respiratory physicians classified the patients from mild to critical type and then cross-checked, a third expert was involved when there was disagreement.

All patients’ general information, days from symptom onset to admission to hospital, from symptom onset to developing into severe/critical type, from severe type to critical type, from symptom onset to death, from symptom onset to discharge, from exposure onset to discharge were included. The median CRP value, positive rate, and distribution of median over time were studied in different course with various initial time, and different periods, including after disease onset, exposure onset, 4 days before and after developing into severe and critical type, and before death.

### 2.3 CRP EXAMINATION

The CRP test was conducted by the Laboratory Department of Xiangyang No.1 People’s Hospital, using Turbidimetric inhibition immune-assay. The reagent was C-reactive protein test kit (Abbott Laboratories), the test instrument is automatic biochemical immunoassay analyzer (Abbott Laboratories ARCHITECT c16200), the normal range of CRP value was 0.00-8.00 mg/L.

### 2.4 STATISCAL ANALYSIS

All statistical analyses were performed using SPSS 20.0. Binary data were described using frequency and percentage. The normality of continuous data was checked. Mean and standard deviation were used to describe variables with normal distribution; otherwise, median (interquartile, IQR) was used. Categorical data were described as frequency (%); the chi-square test was applied to assess significance between groups. All graphs were processed using GraphPad Prism 8.0 and Photoshop CC 14.2 software.

## 3 Results

This study included all of the suspected and laboratory-confirmed 542 patients till Feb 28, 2020. Among the 542 cases, the nucleic acid tests in 142 cases were positive but 9 cases that have data stored in other hospitals cannot be traced and 2 were infants. Therefore, 131 cases were included in the study finally.

### 3.1 GENERAL INFORMATION

Among the included 131 cases, 121 cases were discharged from the hospital while 10 cases died. There were 63 males and 68 females, and the average age was 50.13±17.13 years old. The time from exposure to symptom onset was 9.88±5.26 days, from symptom onset to hospital admission was 4.54±3.10 days, average hospitalization time was 22.38±8.70 days, from symptom onset to severe type was 8.15±5.29 days, from symptom onset to critical type was 11.83±5.19 days, from severe to critical type was 4.29±5.76 days, from symptom onset to death was 18.30±9.65 days, and from symptom onset to discharge from the hospital was 26.87±9.19 days.

### 3.2 CRP RESULTS

The 131 cases underwent 37 laboratory indicators contained 24052 tests in outpatient and hospitalization. This study included all of the CPP results 724 times of tests totally, account for 3.01% of the all results of the indicators.

### 3.3 THE FEATURES OF CRP FROM SYMPTOM ONSET

The 131 patients underwent 724 tests of CRP from the symptom onset to the day 51, the median value was 9.52(2.63-34.10) mg/L. 53.18% of the results exceeded normal range. During the first 30 days, there were 661 tests and the median was 11.69(2.87-39.63) mg/L, among which 55.67% tests were higher than the upper limit of normal value (ULN). In T1-T8, 58.54%, 69.77%, 66.40%, 49.62%, 41.96%,42.68%, 33.33%, and 15.79% of the tests were abnormal, respectively. According to the changing trend plot per day, the CRP value increased during day to day 11, decreased during day 11 to day 17, and then fluctuated around 8.00 mg/L during day 17 to day 30. According to the changing trend plot per 5 days, the CRP value increased from T1 to T3, and decreased from T3 to T6. In T4, the CRP value went back to the normal range. The abnormal period was T1-T3 (Table 1, Fig. 1).

**Table 1.**
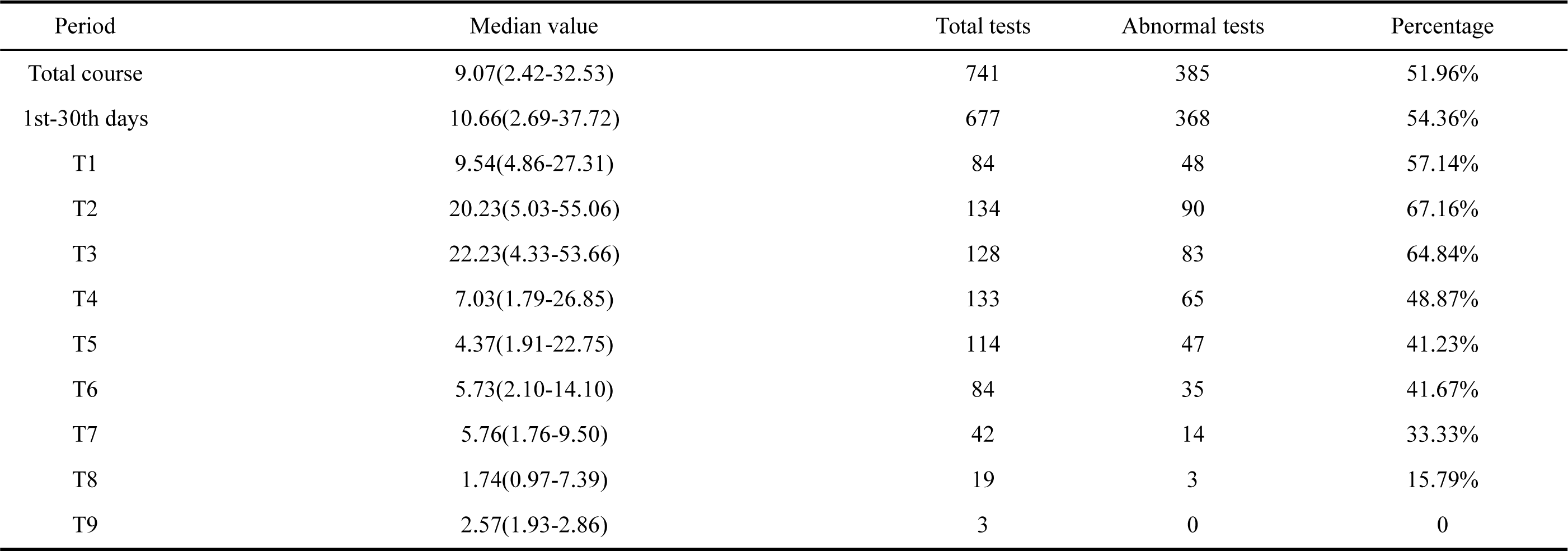
Median and abnormal value in 133 patients

**Fig. 1.**
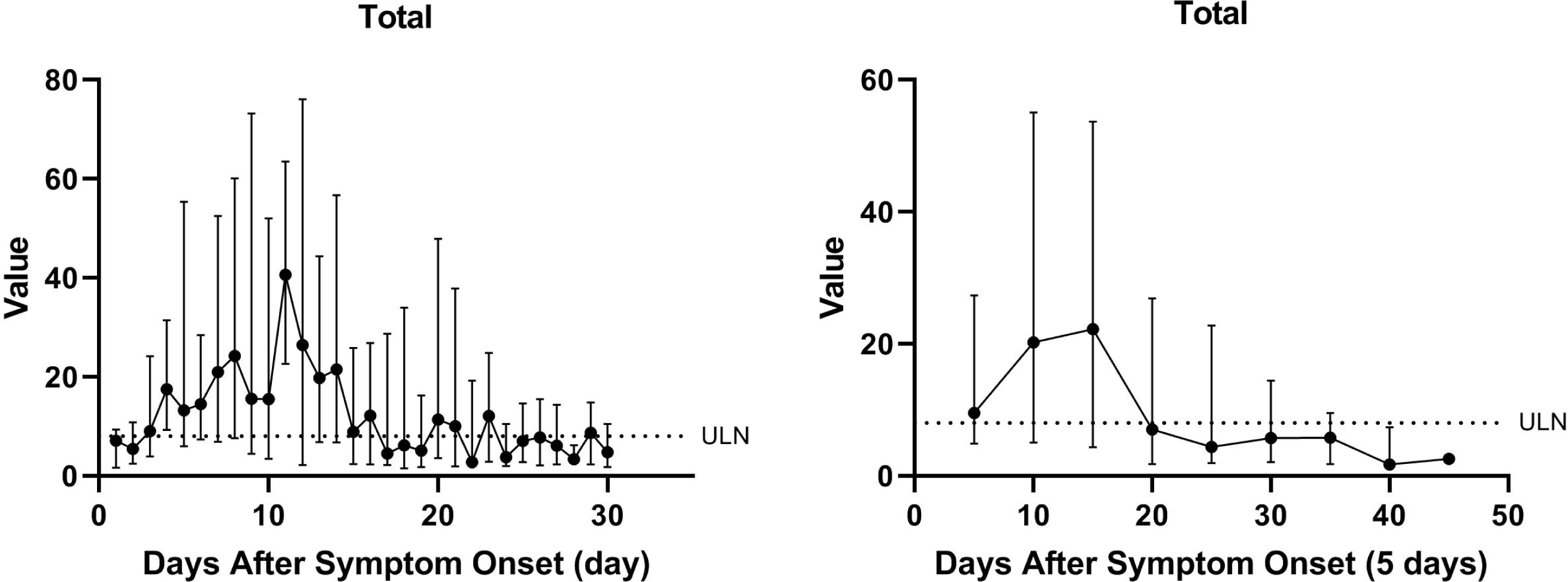
The distribution over time of CRP median in total cohort from symptom onset.

### 3.4 THE FEATURES OF CRP IN 42 CASES FROM EXPOSURE ONSET

We followed up all of the cases and finally defined the 42 cases with a definite exposed date. 42 cases underwent 259 tests during day 1 to day 50, and the median was 15.83(4.08-44.61) mg/L, among which 64.86% of the test result were higher than the ULN. Time from exposure onset to admission was 14.05±5.93 days and from exposure onset to discharge was 36.64±10.10 days. The changes of CRP median from exposure onset indicated that the first median value on the day 8 was 39.08(16.69-46.06) mg/L with the 8 days as a cycle. It fluctuated from exposure onset up until the day 28 accompanying with the peak value around 39.00 mg/L, which presented a cycled-fluctuating tendency and finally decreased during the day 32 to day 50. The abnormal period was day 8 to day 38 and day 48 (Fig. 2).

**Fig. 2.**
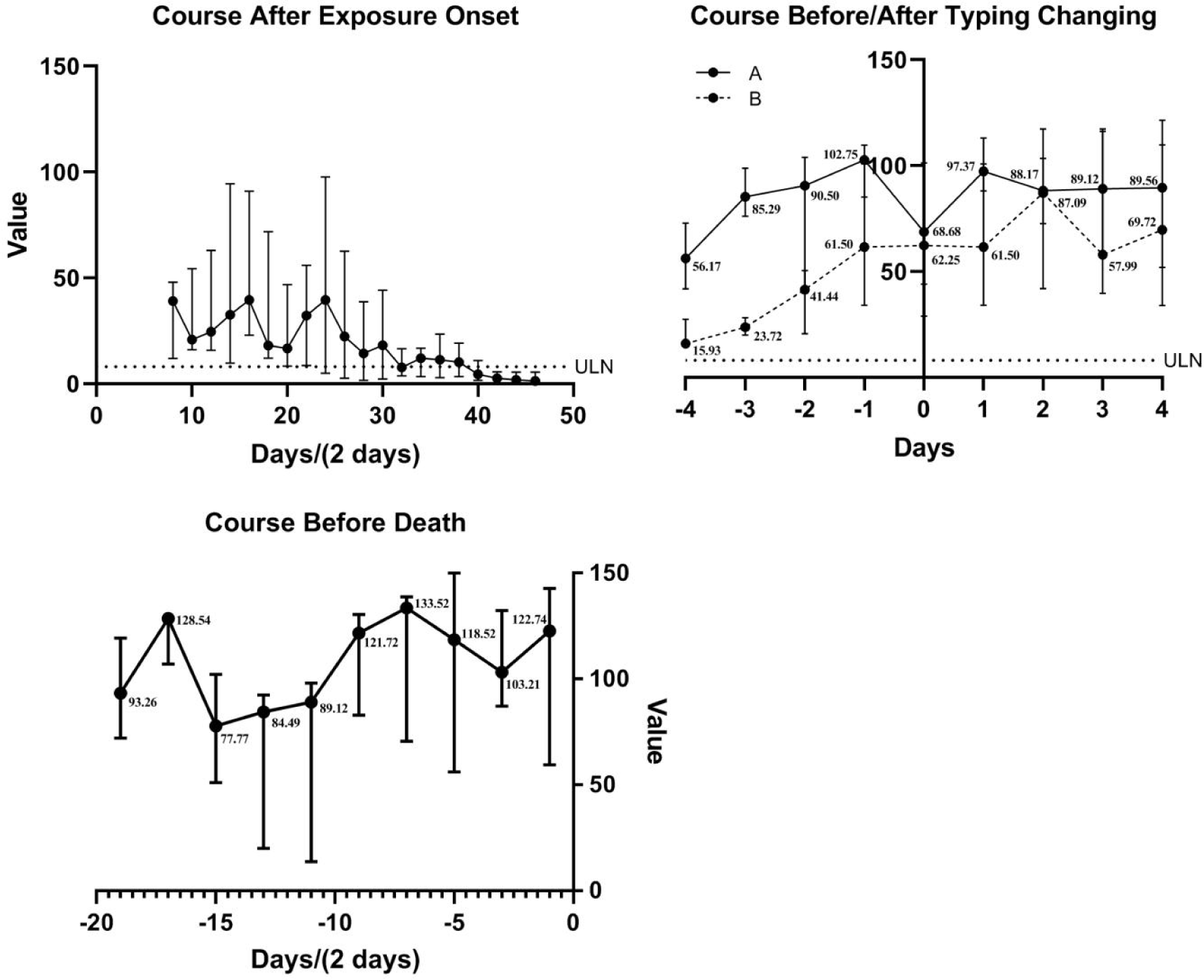
The distribution over time of CRP median during the period from exposure onset, before and after transformation of severe or critical, and before death.

### 3.5 THE FEATURES OF CRP BEFORE AND AFTER TRANSFORMATION OF SEVERE AND CRITICAL TYPE

The transforming date of developing into severe type was set as “0” in 21 severe cases and 18 critical cases, the changes that 4 days before and after “0” was plotted in 21 severe and 18 critical cases. Above the 93 tests, the median was 45.14(24.58-87.09) mg/L, and 91.40% of them exceed the normal extent. The tendency showed that the CPR value was prone to increase from the day -4 (15.93 mg/L) to the day -2 (41.44 mg/L), and decrease from day -2 to day -1 (18.26 mg/L) then decease from day -1 to day 0 (62.25 mg/L). Similarly, the changing trend 4 days before and after transforming date of developing into critical type from 18 cases was plotted. Above the 62 test results, the median was 90.80(55.86-109.59) mg/L, and 95.16% of them exceed the normal extent. The chart showed that the CRP value was increased from the day -4 (56.17 mg/L) to the day -1 (102.75 mg/L) but decreased during day -1 to day 0 (68.68 mg/L) (Fig. 2).

### 3.6 THE FEATURES OF CRP BEFORE DIED

The 58 tests were found between the day -1 to day -16 with the abnormal rate as 100.00%, with the median value as 103.64(64.87-129.7) mg/L in 10 death. The tendency showed that day -16 (77.77mg/L) to the day -8 (133.52mg/L) was prone to increase, and the peak value was 133.52(70.53-138.8) mg/L. CRP median fluctuated about 103.21-133.52 mg/L and the median was 120.63(79.45-136.12) mg/L during day -1 to day -10, nearly presented a plateau level (Fig. 2).

## 4 Discussion

A total of 542 suspected and confirmed COVID-19 cases were included without choice. Finally, we included all hospitalized 131 SARS-CoV-2 test positive cases. All of the laboratory indexes data were collected, and we found that CRP held a high positive rate and presented obviously increased when illness aggravated. According to the time from symptom onset to discharge from hospital, 26.87±9.19 days, we statistically analyzed the data from the day 1 to day 30.

When studying the course from symptom onset, the median of CRP was 9.52(2.63-34.10) mg/L and 54.18% of the total tests exceeded normal range. As shown in the changes per day, the median value of CRP on the day 3 was 9.01 mg/L, which was higher than the normal extent, while the average time from symptom onset to admitted was 4.54±3.10 days, indicating that CRP might have already increased abnormally before admitted. Systematic inflammatory reaction happened with the sign as increased CRP 3 days after symptom onset (12). Therefore, CRP might have potential value for the diagnosis at the early stage. Moreover, it showed that the peak value was 40.63 mg/L on the day 11, which was in consistent with the time from symptom onset to developing into the critical type, and the time of the peak value of the CT score in our cohort (supplement figure). The result indicated that the peak of an inflammatory response, the peak of disease evolvement, and the peak of progress in focus of lungs were synchronous. CRP could reflected the changes of lesion in focus and illness to some extent. Lastly, abnormal tests were distributed within day 3 to day 16, day 20 to day 21 and day 23, meanwhile major symptoms disappeared within the first 2 weeks, and most of the symptoms vanished up to 3 weeks, which was in consistent with the abnormal distribution interval of CRP. These results were partially similar to the report of Zhong *et al* (9).

According to the CRP changes per 5 days, the abnormal results mainly appeared during T1-T3, and T1-T2 held the widest rise extent the abnormal rate was highest in T2, and the peak value (23.24 mg/L) was located in T3. It indicated that most of the tests and median value of the whole CRP were higher than upper limit value and the abnormal interval mainly situated in T1-T3 (day 3 to day 16). The outliers during day 20 to day 21 and day 23 might because of part of the critical cases accompanied complications.

Virus infection usually induce the increment of CRP, while rarely existed substantial elevation. It was acknowledged that CRP>50.00 mg/L was regarded as a standard to exclude virus infection (15, 16), while the median value was 9.52 mg/L and 19.75% of the tests in this cohort showed CRP>50.00 mg/L, which might be another feature of COVID-19. This analysis revealed that SARS-CoV-2 virus infection could induce severe inflammatory response, and substantially and rapidly increased CRP in the early stage. It is different from the manifestations of other diseases. This feature was helpful for early diagnosis of COVID-19.

The time from symptom onset to death was 18.3±9.65 days, which was relatively dispersed. Thus, the corresponding CRP distribution could not describe the features of changes in CRP before death clearly. According to the changes of CRP before death in this study, median value was 77.77 mg/L on the day -16 and then continuously increased on the basis of substantially increasing with the peak value as 133.52 mg/L on the day -8. it maintained about 120.00mg/L 10 days before death,. It indicated that a long time and dramatically elevating up to 120.00 mg/L in CRP might be a characteristic distribution in CRP before died. And revealed this manifestation might induce severe tissue injury and even death. The described distribution showed CRP changes before death accurately and was helpful for us to analyze and estimate the illness.

Due to the differences in the speed of disease progressed, the time for patients to transform from moderate type to severe and critical type was quite different. According to the results, CRP decreased from day -2 to day -1 before moderate type developing into severe type but increased from day -1 to day 0, which was consistent with our findings in symptom study that the symptoms ms appeared to reduction just before the transformation of aggravated clinical type and patients also feel better at that time. Conversely, CRP continuously increased before progressing into critical type, but decreased from day -1 to day 0, and then increased after transformation. However, the scope of focus didn’t decrease in CT (Supplement Figure), which might due to the different time sequences of tissue injury, inflammatory response, symptom, focis absorption, and repair after infection. Overall, CRP caused several inflammatory response-symptoms, and the clinical type was divided depending on lung function, while the peak of inflammation emerged before the peak of dysfunction. This indicated that, the stabilization or decreasing for a while in CRP didn’t represent the improvement of illness, size of lung focus in CT images and pulmonary functions should be concerned closely.

The time of symptom onset always regarded as the disease onset, while virus replication in the infected subject before symptom onset induced tissue damage and inflammatory response. Symptom onset just when the viral load and damage reached a certain extent, thus the complete course should be recorded from exposure onset. As shown in Fig. 3, abnormal median distributed within day 8 to day 30, day 34 to day 38 and day 48, the first test in the day 8, and the peak separately located in the day 8 (39.06 mg/L), day 16 (39.62 mg/L), and day 24 (39.51 mg/L) a cycled-fluctuating tendency. Notably, each peak value was approximate with an interval of 8 days and 3 obvious cycles containing 28 days. The time from exposure to symptom onset was 9.88±5.26 days. The CRP values were at an abnormal peak when first tested on the day 8 since exposure onset, suggesting that the virus has induced inflammatory response with an extensive increase of CRP before symptom onset. This feature was different from other viral infectious diseases and might be a diagnosis index for COVID-19 in the early stage. Additionally, the result indicated a feature of cycled-increased but whether this feature was correlated with the viral replicative cycle still needs further study.

The cases included in our cohort were obtained from single hospital’s inpatient cases, and the evidence value was limited. We analyze the data in outpatient and hospitalization that cannot reflect the features post-discharge. Next, the limited sample size influenced the analysis of CRP data per day. Multi-centered and more samples are needed to verify the reference standard of CRP in severity classification.

The CRP median value of overall tests and most of the results were abnormal with the manifestations of increasing during the early to middle stage. Dramatically increased CRP before symptom onset might be a characteristic of COVID-19 patients. Distribution of CRP from exposure onset showed that the first test was situated at peak and presented as a 3 cycled-distribution overall, which might be correlated with the cycle of virus replication and inflammatory response. CRP decreased 2 day before moderate type developing into severe type and decreased 1 day before severe type progressing into critical type. CRP maintained over 100.00 mg/L prompted poor prognosis.

## Data Availability

Anyone who wants to obtain the original data of this study with reasonable purposes can contact the correspondent author via email.

## CRediT authorship contribution statement

Concept and design: Yunfu Wang, Bin Pei; Recruited patients: Hui Yu, Meiling Zhang, Lei Chen, Haoming Zhu; Data extraction: Guoxin Huang, Gaojing Qu, Meiling Zhang; Performed data analysis: Guoxin Huang, Gaojing Qu; Drafted and revised the manuscript: Hui Yu, Bin Pei; All authors provided critical review of the manuscript and approved the final draft for publication. Guoxin Huang, Gaojing Qu Contributed equally.

## Abbreviations

COVID-19: Coronavirus Disease 2019
CRP: C-reactive Protein
WBC: White Blood Cell
LYMPH: Lymphocyte
ULN: Upper Limit of Normal

## Appendices

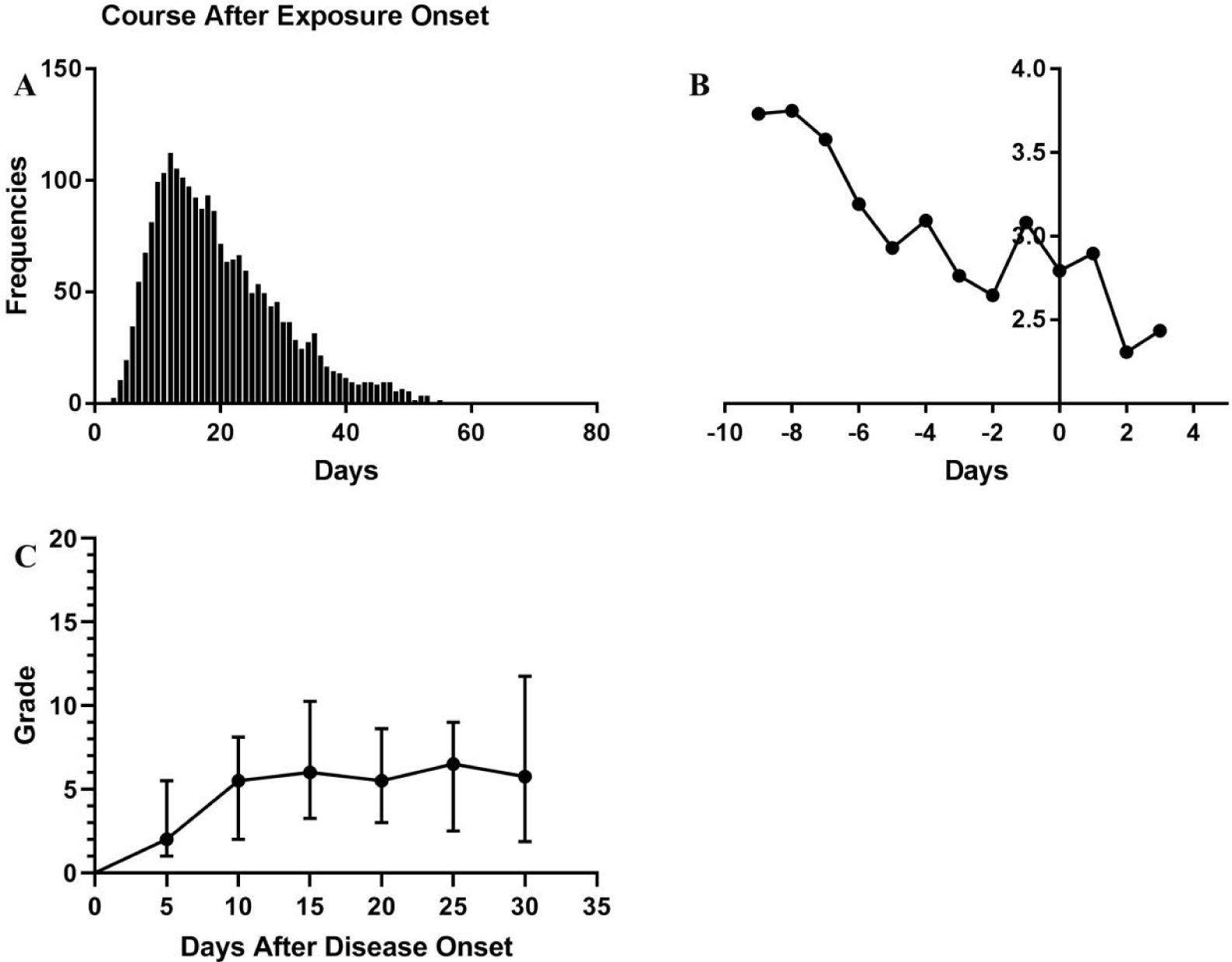
A: The symptoms frequencies over time from exposure onset; B: The frequencies/cases of symptom over time before and after transformation of severe type; C: The CT score over time from symptom onset.

